# Identification of a super-spreading chain of transmission associated with COVID-19

**DOI:** 10.1101/2020.03.19.20026245

**Authors:** Ke Hu, Yang Zhao, Mengmei Wang, Qiqi Zeng, Xiaorui Wang, Ming Wang, Zhishui Zheng, Xiaochen Li, Yunting Zhang, Tao Wang, Shaolin Zeng, Yan Jiang, Dan Liu, Wenzhen Yu, Fang Hu, Hongyu Qin, Jingcan Hao, Jian Yuan, Rui Shang, Meng Jiang, Xi Ding, Binghong Zhang, Bingyin Shi, Chengsheng Zhang

**Author notes:** Corresponding author **Corresponding authors**: Ke Hu, M.D., Department of Respiratory Disease, Renmin Hospital of Wuhan University, Wuhan, China.; Chengsheng Zhang, M.D., Ph.D., Precision Medicine Center, The First Affiliated Hospital of Xi’an Jiaotong University, Xi’an, China. **Author’s contribution:** K.H., Y.Z., M.W., Q.Q.Z., X.R.W., B.H.Z., B.Y.S., C.Z participated in study design; K.H., Q.Q.Z., X.R.W., F.H., H.Y.Q., J.C.H., J.Y., R.S., M.J., X.D., B.Y.S., C.Z. performed data analysis; K.H., Y.Z., M.W., M.W., Z.S.Z., X.C.L., Y.T.Z., T.W., S.L.Z., Y.J., D.L., W.Z.Y., B.H.Z. recruited patients; K.H., Y.Z., M.W., Q.Q.Z., X.R.W., F.H., H.Y.Q., B.Y.S., C. Z. drafted the manuscript; K.H. and C.Z. were responsible for study conception; all authors provided critical review of the manuscript and approved the final draft for publication.

## Abstract

**Background:** Super-spreading events were associated with the outbreaks of SARS and MERS, but their association with the outbreak of COVID-19 remains unknown. Here, we report a super-spreading transmission chain of SARS-CoV-2 involving an index patient, seven cancer patients, 40 health care workers and four family members.

**Methods:** We conducted a retrospective study to identify the index patient and the exposed individuals linked to a chain of transmission associated with COVID-19. We collected and analyzed the data on demographic features, exposure history, clinical presentation, laboratory investigation, radiological examination, and disease outcome of these patients.

**Results:** We identified the index patient and another presumptive “super-spreader”, who initiated and amplified a super-spreading transmission chain associated with COVID-19, respectively. There were 31 female and 21 male patients in this cohort, and the median age was 37 years (range: 22-79 years). Each of them had an exposure history with the index patient or his close contacts. Approximately 87% (45/52) of the patients had fever or other symptoms, 96% (50/52) had abnormal chest CT-scan findings, 86% of the tested patients (39/45) were positive for SARS-CoV-2 in the nasopharyngeal or throat swab specimen, 85% of the tested patients (29/34) were positive for SARS-CoV-2-specific IgM and/or IgG, 15% of the RT-PCR positive patients were tested negative for the specific IgM and/or IgG at the convalescent phase, and 15% of the RT-PCR negative patients were tested positive for the specific IgM and/or IgG. The severe patients experienced a significant decrease in oximetry saturation, lymphocyte, and platelet counts, along with a significant increase in C-reactive protein, D-dimer, and lactate dehydrogenase. All six fatal cases had comorbidities and five of the seven cancer patients (71%) died within 2-20 days of the disease onset.

**Conclusions:** The super-spreading events were associated with the outbreak of COVID-19 in Wuhan and its impact on disease transmission warrants further investigation. Cancer patients appeared highly vulnerable to COVID-19. The finding that a significant portion of SARS-CoV-2 infected patients were tested negative for the serum specific IgM and IgG at the convalescent phase should be addressed by additional studies.

## INTRODUCTIONS

The outbreak of coronavirus disease 2019 (COVID-19), which was caused by the novel coronavirus SARS-CoV-2, has posed tremendous challenges to the international communities^1-8^. As of March 18, 2020, there were 81,116 confirmed cases and 3231 deaths in China^9^; Globally, 160 counties and territories have reported cases of COVID-19, including 191,127 confirmed cases and 7807 deaths^9^. In response to “the alarming levels of spread and severity”, the World Health Organization (WHO) has characterized COVID-19 as a pandemic^10^. This is the first pandemic caused by a coronavirus. Coronaviruses (CoV) are a large family of RNA viruses that cause a variety of mild and severe diseases in humans and animals^11^. Prior to the COVID-19, there were two severe outbreaks of human coronavirus diseases in the past two decades, the severe acute respiratory syndrome (SARS) and the Middle East respiratory syndrome (MERS)^12-16^, which were caused by SARS-CoV and MERS-CoV, respectively. SARS was first reported in November 2002 in Guangdong Province, China. There were 8,422 confirmed SARS cases, including 919 deaths in 32 countries between November 2002 and August 2003^17^. MERS was first identified in September 2012 in Saudi Arabia. There were 2494 confirmed MERS cases, including 858 deaths in 27 countries between September 2012 and November 2019^18^. Super-spreading events, by which an individual patient spread an infection to a large number of susceptible people, were associated with the outbreaks of SARS and MERS^19-22^. While case clusters with human-to-human transmission of SARS-CoV-2 have been reported^3-5,23-25^, it remains unknown whether super-spreading events occurred in the outbreak of COVID-19. Here, we conducted a retrospective study and identified a super-spreading chain of transmission associated with COVID-19.

## METHODS

### Patients and data collection

This retrospective study was approved by the institutional review board of Renmin Hospital of Wuhan University (No. WDRY2020-K019). Oral consent was obtained from the patients or their family members, whereas written informed consent was waived by the Provincial and National Health Commissions in China under the exceptional circumstances for investigation of an ongoing disease outbreak. In this cohort, we identified a super-spreading chain of transmission involving 52 linked patients with COVID-19 in three separate hospitals in Wuhan, the epicenter of an outbreak of COVID-19 across China. The data we retrieved from the electronic medical records included demographic features, comorbidities, clinical presentation, laboratory investigation, RT-PCR testing for SARS-CoV-2, serum specific IgM and IgG antibodies, chest computed tomographic (CT) scan, and the disease outcome. The contact and exposure history were also collected through communications with the patients and/or their family members.

### Case definition

All patients with COVID-19 enrolled in this study were diagnosed and classified according to World Health Organization interim guidance and the “Guidelines on the Diagnosis and Treatment of the Novel Coronavirus Infected Pneumonia” developed by the National Health Commission of People’s Republic of China^26,27^. The severe cases showed at least one of the following presentations: (1) Respiratory distress, Respiratory rate (RR)⩾30 times/min; (2) At rest, oxygen saturation⩽93%; (3) Arterial partial pressure of oxygen (PaO_2_) /Fraction of Inspired Oxygen (FiO_2_) 300mmHg (1mmHg=0.133kPa). Patients who had close contact with the index patient were defined as secondary cases. Patients who contracted the disease from the secondary cases were classified as tertiary cases.

### Laboratory investigations

The routine laboratory investigations were performed by the certified clinical diagnostic laboratories inside the designated hospitals. The RT-PCR testing for SARS-CoV-2 were conducted by the Standard Operation Procedures published previously^28,29^. Serum specific IgM and IgG antibodies against SARS-CoV-2 were detected using the immunoassay kit provided by the YHLO Biotech Co., Ltd. (Shenzhen, China) and the fully-automated chemiluminescence immunoassay analyzer (UniCel DxI800, Beckman Coulter, Inc., USA) according to the instructions of the manufacturers. The immune activities were measured as a relative light unit (RLU). The analyzer automatically converts the RLU from the immunoassay into an absolute unit by fitting the standard master calibration curve. A cutoff value of ⩾10.0 AU/ml is considered as positive for both antibodies.

### Statistical analysis

For statistical analysis, all of the continuous variables were performed by Shaprio-wilk test and Levene test to analyze normality and homogeneity. Continuous variables were expressed as median (IQR) and compared with T test or Mann-Whitney U test. Categorical variables were expressed as number (n/N%) and compared by χ^2^ test, continuity-adjusted χ^2^ test or Fisher’s exact test. A two-sided α of less than 0.05 was considered statistically significant. Statistical analyses were done using the SPSS 22.0 software.

## RESULTS

In this report, we conducted a retrospective study and identified a super-spreading chain of transmission involving the index patient, seven cancer patients, 40 health care workers and four family members (Figure 1 and Table 1).

**Table 1.**
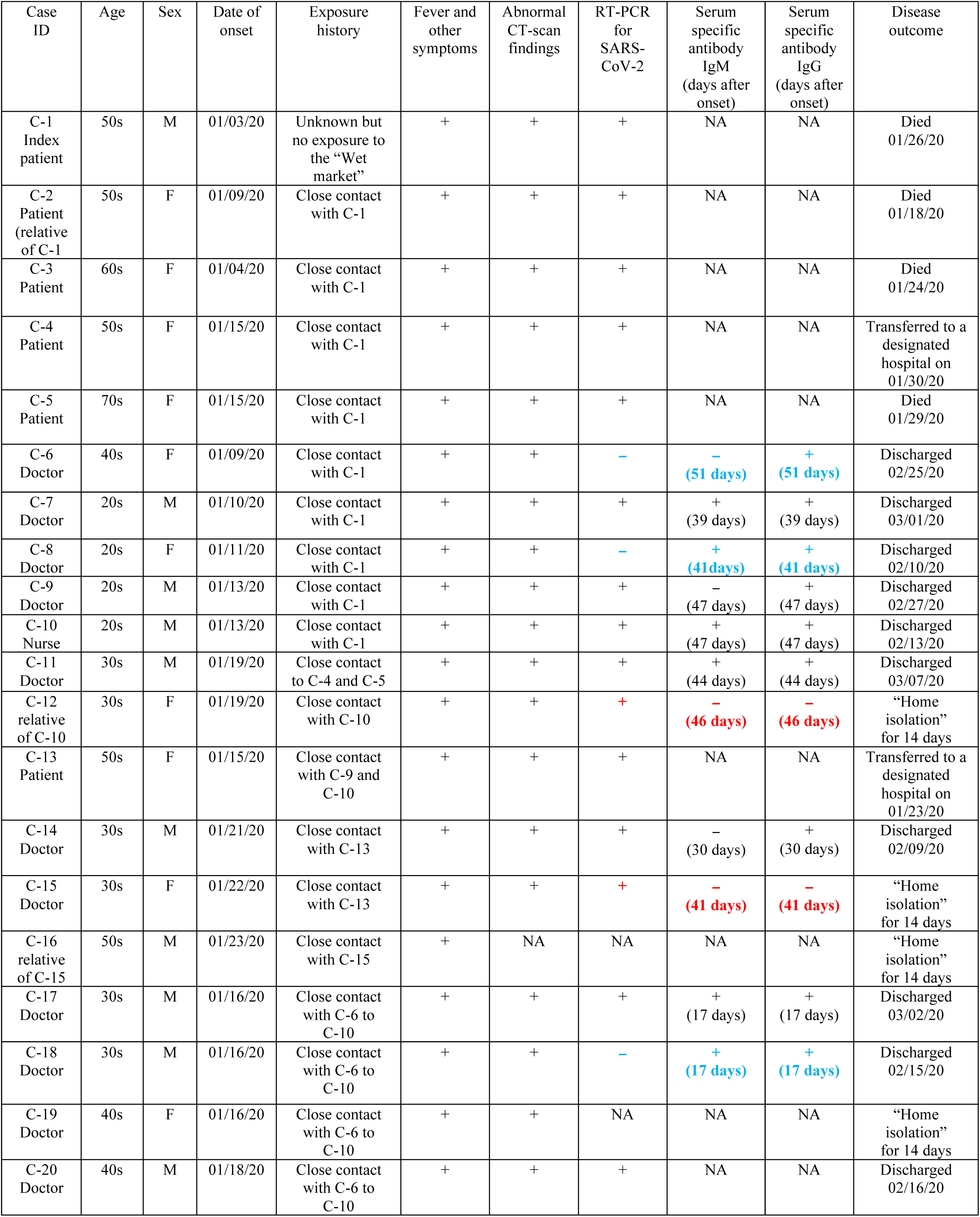
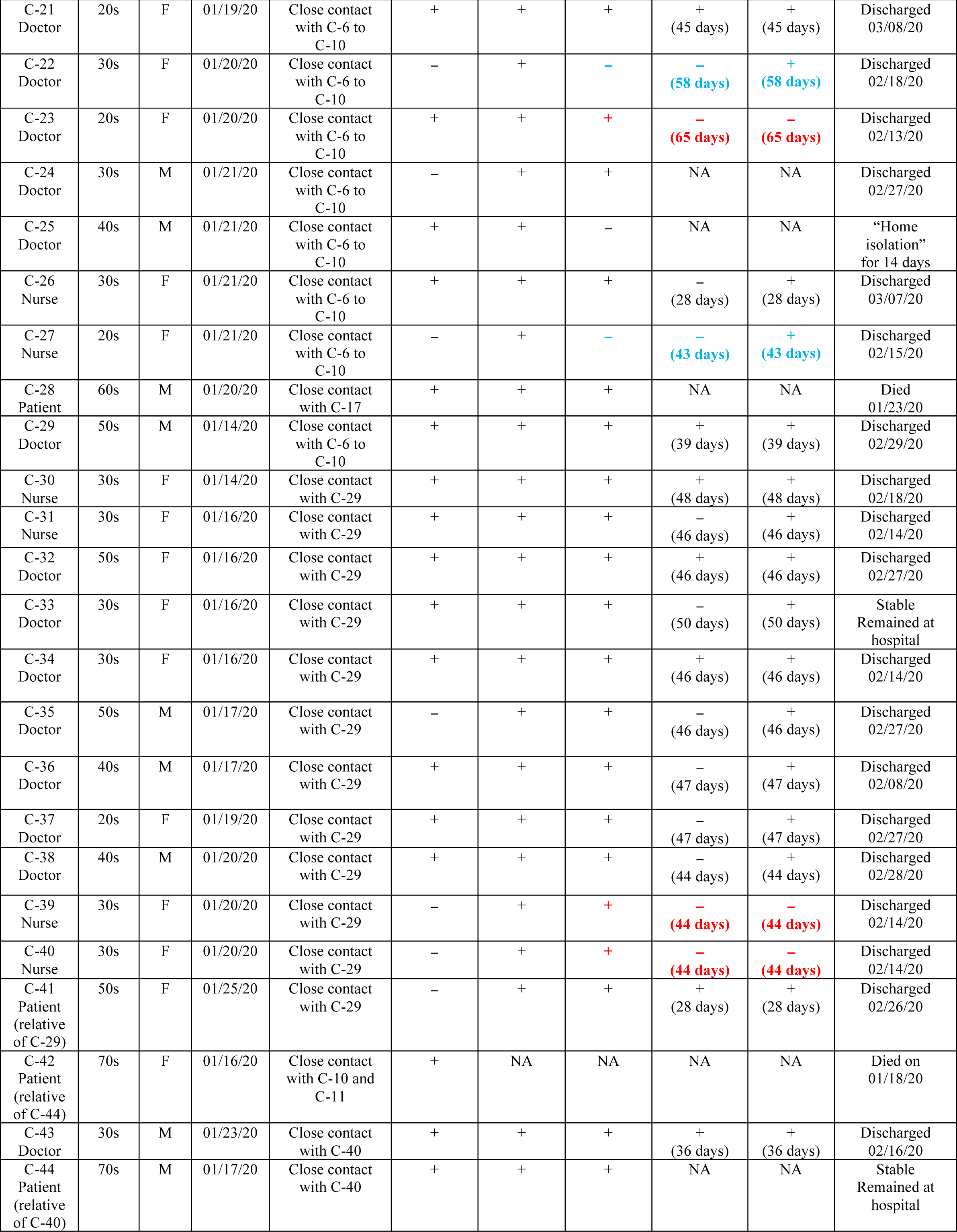
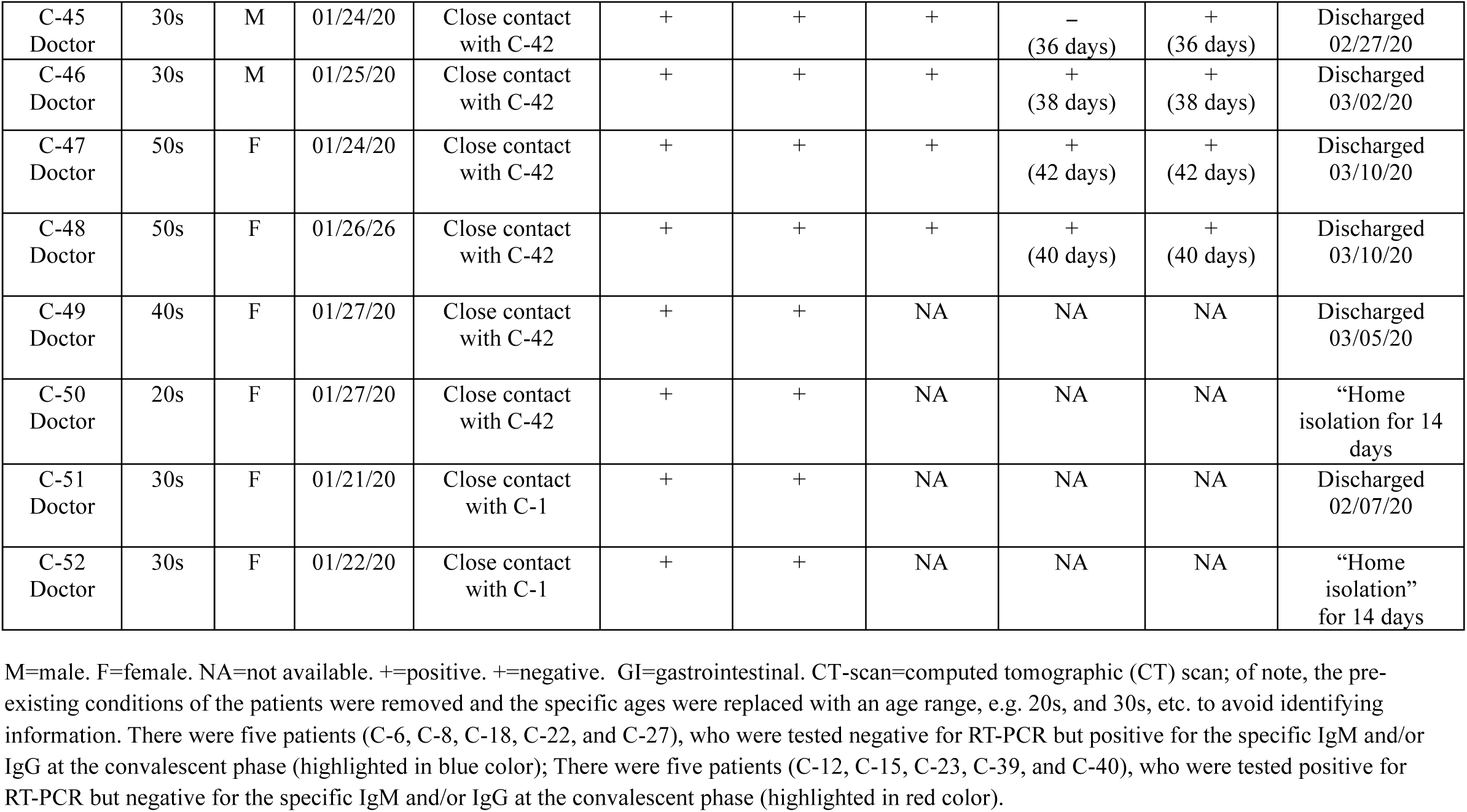
Demographics, clinical features and laboratory findings of patients associated with COVID-19.

**Figure 1.**
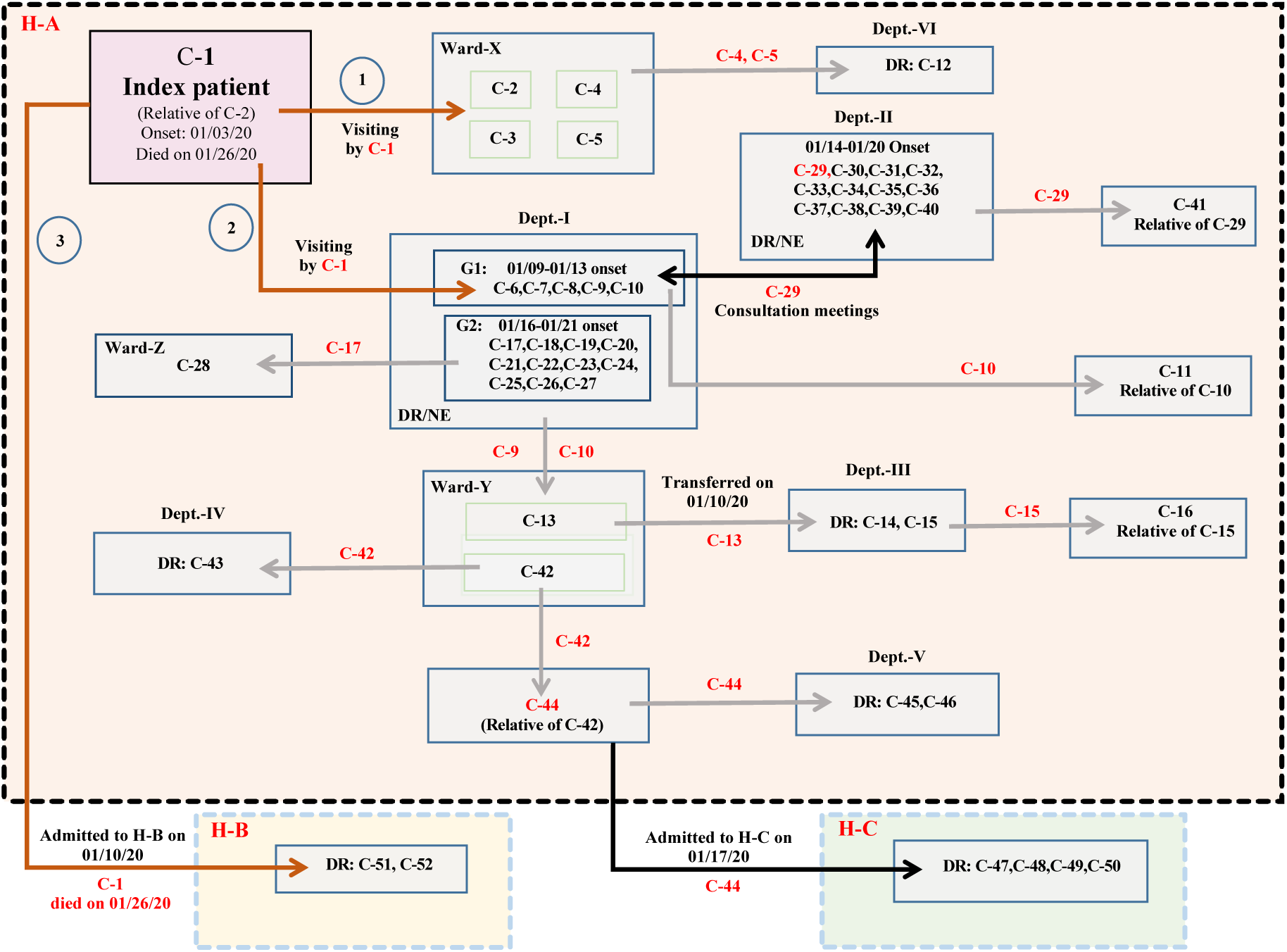
A schematic diagram showing a super-spreading chain of transmission associated with COVID-19. There were 52 individuals in this cohort at multiple clinical departments in three separate hospitals in Wuhan, China. Dept.=Department; DR=Doctor; G1=Group 1; G2=Group 2; GI=Gastrointestinal; H-A=Hospital A; H-B=Hospital B; H-C=Hospital C; NE=Nurse.

### Identification of the index patient (C-1)

On January 3, 2020, a male patient in his 50s developed fever (38.8°C), headache, chest pain and myalgias when he visited his relative in a ward room (Ward-X in Figure 1) shared by four female cancer patients at Hospital A. He initially thought that he had a “cold” and continued to visit this ward until he was admitted to hospital B on January 10, 2020. His Chest CT-scan findings showed a rapid progression of bilateral infiltrates in the lungs (Figure 2). He received antibiotics treatment for one week but was ineffective. On January 18, he was tested positive for SARS-CoV-2 and diagnosed with COVID-19. He died of severe respiratory failure on January 26, 2020. While the exact exposure history of this patient remained unclear, he did not visit the “Huanan Wet Market”, the suspected origin for this outbreak in Wuhan. Of note, the “Huanan Wet Market” is located in the Hankou District of Wuhan city, which is separated from the Wuchang District by the Yangtse River. The two hospitals (A and B) where the index patient visited were located in Wuchang District.

**Figure 2.**
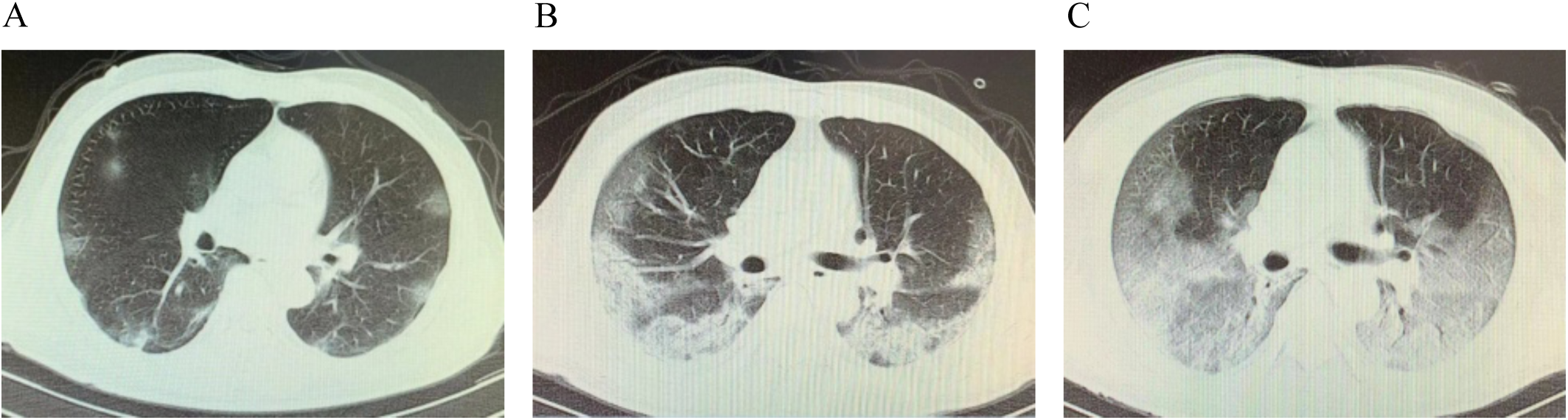
Representative chest CT images of the index patient (C-1) showing rapid progression of bilateral infiltrating shadows in the lungs. A. An image taken on 01/10/2020 (the admission day); B. An image taken on 01/13/2020 (three days after admission); C. An image taken on 01/16/2020 (6 days after admission).

### A super-spreading event initiated by the index patient

Eleven people presented fever and abnormal CT-scan findings after close contacts with the index patient C-1 between January 3 and January 10, 2020 (Figure 1 and Table 1). Below was a timeline for the initial transmission events. (1) C-1 visited his relative C-2 in Ward-X that was shared by other three cancer patients (C-3, C4, and C-5) at the Department-I of hospital A when he first developed fever and other symptoms on January 3. All four cancer patients had close contacts with C-1 and subsequently developed fever (38°C-40°C) and abnormal CT-scan findings between January 4 and January 15. All were RT-PCR positive for SARS-CoV-2, but the assays for the serum specific antibodies were not available at that time. All four patients did not have other exposure records and epidemiological history before their disease onsets. Three of them died of severe respiratory failure within 9-20 days after the onsets. C-2 developed symptoms on January 9 (6 days after the onset of C-1) and died on January 17. C-3 developed symptoms on January 4 (one day after her close contact with C-1) and died on January 24. C-4 developed symptoms on January 15 and transferred to a designated hospital on January 30, where she remained at a stable condition after the treatment. C-5 developed symptoms on January 15 and died on January 29.

On January 3, C-1 also visited the office area of the health care workers at the Department-I of hospital A to discuss the treatment plan for his relative C-2. Four doctors (C-6, C-7, C-8 and C-9) and one nurse (C-10) developed fever and abnormal CT-scan findings between January 9 and January13 (within 10 days after close contacts with C-1). In addition, a cluster of eleven health care workers (C-17 to C-27) from the same department who had close contact with their co-workers (C-6 to C-10) presented fever and abnormal CT-scan findings between January 16 and January 21 (within 3-12 days after their exposure to the secondary cases). All the health care workers were previously healthy and did not have other exposure records and epidemiological history (Figure 1 and Table 1).

On January 10, C-1 was admitted to hospital B, where he was diagnosed with COVID-19. Two doctors (C-51 and C-52) had close contacts with C-1 when they met with him on a medical consultation meeting to discuss his treatment plan. They presented fever and abnormal CT-scan findings on January 21 and January 26, respectively. Both of them were previously healthy and did not have other exposure records and epidemiological history. Our findings suggested that C-1 was the source patient and considered as a “super-spreader” who initiated this super-spreading chain of transmission.

### Another super-spreading event initiated by a doctor from Department-II in hospital A

Between January 14 and January 20, 2020, a total of twelve doctors and nurses (C-29 to C-40) from Department-II in hospital A developed fever and abnormal CT-scan findings (Figure 1 and Table 1). All twelve individuals were previously healthy, and none of them except C-29 had other exposure records and epidemiological history. C-29, a male doctor in his 50s, attended medical consultation meetings between January 3 and January 8 at the Department-I to discuss treatment plans for two lung cancer patients (C-2 and C-3). Of note, Department-I and Department-II were located at two separate buildings in hospital A. C-29 had close contact with the health care workers (C-6 to C-10) on the consultation meetings before he developed fever and abnormal CT-scan findings, and was diagnosed with COVID-19 on January 14. All eleven co-workers (C-30 to C-40) had close contacts with C-29 before his disease onset. In addition, C-29’s relative (C-41) was diagnosed with COVID-19 on January 25. Our findings indicated that C-29 was likely the source patient who was responsible for introduction of COVID-19 into this cluster, and therefore was considered as a “super-spreader”.

### A cluster of tertiary cases linked to a gastric cancer patient

C-13, a female with gastric cancer, was initially hospitalized in Ward-Y at Department-I in hospital A. She was transferred to Department-III on January 10 and had surgical operation on January 12. She had close contacts with C-9 and C-10 who were in charge of her treatment at Department-I, However, she did not present fever and other symptoms until January 15 (i.e. 3 days after her surgical operation). She was transferred to a designated hospital after her confirmation with COVID-19 on January 23 and remained at a stable condition. Two Doctors (C-14 and C-15) who conducted the operation procedures were diagnosed with COVID-19 on January 21 and January 22, respectively. In addition, C-16 (C-15’s relative) showed fever and other symptoms on January 23 after close contact with C-16 at home.

### A cluster of tertiary cases linked to a colon cancer patient

C-42, a female colon cancer patient, was admitted to the same hospital ward room (Ward-Y) as C-13 on January 10 and treated by the same medical team (C-9 and C-10). She developed fever (39.9°C) and other symptoms on January 16, and died of severe respiratory failure on January 18. She did not take CT-scan and RT-PCR testing due to her sudden death. C-43, a male doctor from Department-V examined C-42 on January 15, and was diagnosed with COVID-19 on January 23 (8 days after his close contact with C-42). Moreover, C-44 was accompanying his relative C-42 in hospital A and diagnosed with COVID-19 on January 17 (one day after the disease onset of C-42). Two doctors (C-45 and C-46) who conducted the medical treatment on C-44 at Department-V in hospital A were diagnosed with COVID-19 on January 24 and January 25, respectively. Furthermore, C-44 was transferred to hospital C on January 17. Four doctors (C-47, C-48, C-49, and C-50) at hospital C had close contacts with C-44, and developed fever and other symptoms between January 24 and January 27. Our findings also suggested that C-44 was a potential “super-spreader” since he transmitted the disease to six people who had direct contacts with him.

### Additional tertiary cases

(1) C-11 was diagnosed with COVID-19 after direct contact with his relative C-10 at home. (2) C-12, a male doctor from the Department-VI, conducted medical treatment on C-4 and C-5 on January 14, and was diagnosed with COVID-19 on January 19 (5 days after direct contacts with C-4 and C-5). (3) C-28, a male liver cancer patient in his 60s (Ward-Z), was diagnosed with COVID-19 on January 20 (4 days after the disease onset of his doctor C-17) and died of respiratory failure on January 23, 2020.

### Clinical features and laboratory findings

(Figure 1, Table 1, and Table 2). There were 31 female and 21 male patients in this cohort, and the median age was 37 years (range: 22-79 years). None of them were linked to the “Huanan Wet Market”, the suspected origin for this outbreak in Wuhan. Each of them, however, had a contact history with the index patient C-1 or his close contacts. Approximately 87% (45/52) of the patients had fever or other symptoms, 96% (50/52) had abnormal chest CT-scan findings, 86% of the tested patients (39/45) were positive for SARS-CoV-2 in the nasopharyngeal or throat swab specimen, 85% of the tested patients (29/34) were positive for SARS-CoV-2-specific IgM and/or IgG, 15% of the RT-PCR positive patients were tested negative for the specific IgM and/or IgG at the convalescent phase, and 15% of the RT-PCR negative patients were tested positive for the specific IgM and/or IgG. The severe patients experienced a significant decrease in oximetry saturation, lymphocyte, and platelet counts, along with a significant increase in C-reactive protein, D-dimer, and lactate dehydrogenase. Five of the seven cancer patients (71%) died within 2-20 days of the disease onset. All six fatal cases had comorbidities, whereas all the health care workers were previously heathy individuals.

**Table 2:** Laboratory findings of patients with COVID-19.

|  | All patients |  | Severe cases |  | Non-severe cases |  | P value |
| --- | --- | --- | --- | --- | --- | --- | --- |
|  | N | IQR or n/N | N | IQR or n/N | N | IQR or n/N |  |
| White blood cell count, $\times 10^9/L$ | 44 | 4.26 ( 3.65– 5.94 ) | 9 | 5.87 ( 3.37 - 7.61 ) | 35 | 4.13 ( 3.72 – 5.50 ) | 0.163 |
| <4 |  | 18/44 ( 41% ) |  | 3/9 (33% ) |  | 15/35 (43% ) | 0.89 |
| $\geq 4$ | | 26/44 ( 59% ) | | 6/9 (67% ) | | 20/35 (57% ) | / |
| Neutrophil count, $\times 10^9/L$ | 44 | 2.74( 1.90 – 4.30 ) | 9 | 4.37 ( 1.78 – 6.34 ) | 35 | 2.65 ( 2.03 – 3.79 ) | 0.176 |
| Lymphocyte count, $\times 10^9/L$ | 44 | 0.95 ( 0.66 - 1.56 ) | 9 | 0.30 ( 0.20 – 0.62 ) | 35 | 1.08 ( 0.86 – 1.75 ) | 0.000 |
| <1.0 |  | 24/44 (55% ) |  | 9/9 (100% ) |  | 15/35 ( 43% ) | 0.007 |
| $\geq 1.0$ | | 20/44 ( 45% ) | | 0/9 | | 20/35 ( 57% ) | / |
| Haemoglobin, g/L | 44 | 13.45( 11.50 – 14.50 ) | 9 | 13.30 ( 9.65 – 14.95 ) | 35 | 13.60 ( 12.10 - 14.50 ) | 0.423 |
| Platelet count, $\times 10^9/L$ | 43 | 172.00 (132.00 – 217.00 ) | 9 | 108.00 ( 64.50 - 153.50 ) | 34 | 184.00 ( 158.25 – 225.25 ) | 0.001 |
| <162 |  | 18/43 ( 42% ) |  | 7/9 ( 78% ) |  | 11/34 ( 32% ) | 0.038 |
| $\geq 162$ | | 25/43 ( 58% ) | | 2/9 ( 22% ) | | 23/34 ( 68% ) | / |
| Oximetry saturation, % | 19 | 92.00 ( 88.0 – 98.0 ) | 9 | 88.0 ( 80.0 - 91.0 ) | 10 | 97.5 ( 94.0 - 98.0 ) | 0.006 |
| C-reactive protein, mg/L |  |  |  |  |  |  | 0.001 |
| $\leq 5$ | | 18/36 ( 50% ) | | 0/9 | | 18/27 ( 67% ) | / |
| $> 5$ | | 18/36 ( 50% ) | | 9/9 ( 100% ) | | 9/27 ( 33% ) | / |
| Fibrinogen, g/dL | 27 | 2.89 ( 2.47 - 4.09 ) | 8 | 3.86 ( 2.31 - 4.36 ) | 19 | 2.87( 2.47 – 3.77 ) | 0.192 |
| Prothrombin time, s | 28 | 11.15 ( 10.53 – 11.86 ) | 9 | 10.90 (10.20 - 11.90 ) | 19 | 11.20 ( 10.60 – 11.90 ) | 0.694 |
| Activated partial thromboplastin time, s | 27 | 29.80 ( 27.40 – 32.80 ) | 8 | 29.90 ( 26.10 – 33.25 ) | 19 | 29.40 ( 27.40 – 32.30 ) | 0.777 |
| D-dimer, mg/L | 28 | 0.54 ( 0.20 – 2.01 ) | 9 | 2.35 ( 1.04 – 4.11 ) | 19 | 0.23 ( 0.17 – 0.54 ) | 0.000 |
| Albumin, g/L | 41 | 41.00 ( 36.25 – 42.55 ) | 9 | 33.40 ( 29.10 – 42.55 ) | 32 | 41.00 ( 38.88 – 42.68 ) | 0.053 |
| Alanine aminotransferase, U/L | 41 | 20.00 ( 12.00 – 32.00 ) | 9 | 27.00 ( 20.00 – 33.50 ) | 32 | 18.50 ( 11.25 – 32.50 ) | 0.313 |
| Aspartate aminotransferase, U/L | 41 | 22.00 ( 18.00 - 27.00 ) | 9 | 27.00 ( 21.50 – 40.00 ) | 32 | 21.00 ( 17.25 – 26.00 ) | 0.028 |
| ≤32 |  | 35/41 ( 85% ) |  | 5/9 ( 56% ) |  | 30/32 (94% ) | 0.015 |
| >32 |  | 6/41 ( 15% ) |  | 4/9 ( 44% ) |  | 2/32 ( 6% ) | / |
| Alkaline phosphatase , U/L | 34 | 60.80 ( 51.00 – 76.18 ) | 8 | 94.75 ( 48.68– 143.13 ) | 26 | 59.05 (51.00 – 70.75 ) | 0.18 |
| Bilirubin, mmol/L | 41 | 9.50 ( 7.55 – 13.05 ) | 9 | 13.00( 8.50 – 24.47 ) | 32 | 9.10 (7.33 – 12.08 ) | 0.041 |
| Potassium, mmol/L | 38 | 3.87 (3.63 - 4.18 ) | 8 | 3.50 ( 3.10– 4.13 ) | 30 | 3.93 ( 3.67 – 4.19 ) | 0.082 |
| Urea, mmol/L | 40 | 4.07 ( 3.54 – 5.48 ) | 8 | 7.12 ( 5.52 – 13.85 ) | 32 | 3.85 ( 3.38 – 4.56 ) | 0.003 |
| Sodium, mmol/L | 39 | 139.90 ( 138.20 - 141.10 ) | 9 | 137.30 ( 132.75 - 142.65 ) | 30 | 139.90 ( 138.40 - 141.20 ) | 0.223 |
| Creatinine, μmol/L | 41 | 57.0 ( 48.0 – 75.0 ) | 9 | 72.00 ( 43.00 – 98.30 ) | 32 | 57.00 ( 47.75 – 74.75 ) | 0.25 |
| ≤80 |  | 33/41 ( 80% ) |  | 5/9 ( 56% ) |  | 28/32 ( 88% ) | 0.097 |
| >80 |  | 8/41 ( 20% ) |  | 4/9 ( 44% ) |  | 4/32 ( 12% ) | / |
| Creatine kinase, U/L | 35 | 72.00 ( 34.00 – 101.00 ) | 9 | 28.00 ( 20.00 – 216.50 ) | 26 | 74.50 ( 40.75 – 95.00 ) | 0.406 |
| ≤170 |  | 31/35 ( 89% ) |  | 6/9( 67% ) |  | 25/26 ( 96% ) | 0.044 |
| >170 |  | 4/35 ( 11% ) |  | 3/9 ( 33% ) |  | 1/26 ( 4% ) | / |
| Lactate dehydrogenase, U/L | 35 | 193.0 ( 161.0 – 228.0 ) | 9 | 247.0 ( 202.5 – 361.0 ) | 26 | 184.0 ( 157.25 - 202.0 ) | 0.001 |
| ≤214 |  | 25/35 ( 71% ) |  | 3/9 ( 33% ) |  | 22/26 ( 85% ) | 0.007 |
| >214 |  | 10/35 ( 29% ) |  | 6/9 ( 67% ) |  | 4/26 ( 15% ) | / |

## DISCUSSION

In this study, we identified a super-spreading chain of transmission associated with COVID-19 at the early stage of the outbreak in Wuhan. In addition, we were able to find the index patient C-1 and C-29 as the presumptive “super-spreaders” who initiated and amplified this chain of transmission, respectively. We understand that “super-spreader” is still a vague term without a strict scientific definition, we followed a definition adopted by the epidemiologists during the SARS outbreak, which defined the “super-spreader” as an individual with transmission of SARS to at least eight contacts^19^. Therefore, C-1 and C-29 were considered as the super-spreaders because each of them passed the infection of SARS-CoV-2 on to at least eleven contacts. Since super-spreading events were associated with the outbreaks of SARS and MERS^19-22^, it would be not a surprise to discover super-spreading events associated with COVID-19.

Our study has a number of unique features. (1) By coincidence, this transmission chain started from a hospital ward shared by four female cancer patients, and the female health care workers (total of 23) outnumbered the male co-workers (total of 17). As a result, there were 60% female (31/52) and 40% (21/52) male patients in this cohort, which were different from other reports ^3-5^. (2) we were able to identify several distinct clusters of cases based on the exposed time and the date of disease onset, which helped connecting all the related activities and events. (3) There were seven cancer patients in this study, and five of them (71%) died of severe respiratory failure within 2-20 days of their disease onsets, suggesting that cancer patients were highly vulnerable to COVID-19 and had rapid disease progression and high mortality rate due to immune suppression and comorbidities. (4) Approximately 13.5% (7/52) of patients were asymptomatic individuals in this study (Table 1), it would be important to understand the potential contributions of these individuals to the disease transmission if similar portion of asymptomatic patients existed in the general populations with COVID-19^25^. (5) Approximately 15% of the RT-PCR negative patients were tested positive for the specific IgM and IgG. On the other hand, 15% of the RT-PCR positive patients were tested negative for the specific IgM and/or IgG at the convalescent phase. The discrepancy between the results of the RT-PCR assay and serological testing suggested the existence of possible “false negative” and/or “false positive” with these assays. Both scenarios may significantly affect the clinical diagnosis and should be addressed accordingly by additional studies given the fact that strict clinical validation studies were not performed on any of these assays due to the urgent need for screening and diagnosis of COVID-19 during the disease outbreak. In particular, it would be a huge concern if a significant portion of SARS-CoV-2 infected patients were tested negative for the specific IgM and IgG at the convalescent phase.

A large number of health care workers were infected with SARS-CoV-2 in our cohort, which were consistent with a report that 41% of the 138 hospitalized patients at one of the major tertiary hospitals in Wuhan were hospital-related transmission^30^. These findings suggested that serious nosocomial transmission occurred in the early stage of this outbreak, which likely changed the dynamics of COVID-19 and contributed significantly to the widespread of the disease in a short period of time in Wuhan and other neighboring cities in Hubei Province.

We recognized that our study had a number of limitations and the findings need to be interpreted with caution. (1) Some cases had incomplete records of the exact exposure time and epidemiological history. (2) RT-PCR and serological testing were not carried out for some cases due to the irregular service during this outbreak. (3) The virus genomes of SARS-CoV-2 in this cohort were not were sequenced.

In conclusion, we identified a super-spreading chain of nosocomial transmission that occurred in the outbreak of COVID-19 in Wuhan. Given the fact that COVID-19 has been spreading rapidly in many other countries worldwide, it is conceivable that more super-spreading events will be identified in near future, and its impact on the disease outbreak and transmission warrant further investigation. The finding that a significant portion of SARS-CoV-2 infected patients were tested negative for the specific IgM and IgG at the convalescence stage should be addressed by additional studies.

## Data Availability

The data is available upon request.

## Acknowledgment

We are extremely grateful to all the patients and their families for participation of this study and providing all the valuable information. We also thank our colleagues for their kind help and strong support.

## References

1. Zhu N, Zhang D, Wang W, et al. A novel coronavirus from patients with pneumonia in China, 2019. N Engl J Med 2020; 382:727–733.

2. Lu R, Zhao X, Li J, et al. Genomic characterisation and epidemiology of 2019 novel coronavirus: implications for virus origins and receptor binding. Lancet 2020; 395(10224): 565–574.

3. Huang C, Wang Y, Li X, et al. Clinical features of patients infected with 2019 novel coronavirus in Wuhan, China. Lancet 2020; 395(10223): 497–506.

4. Li Q, Guan X, Wu P, et al. Early transmission dynamics in Wuhan, China, of novel coronavirus–infected pneumonia. N Engl J Med 2020. DOI: 10.1056/NEJMoa2001316

5. Chen N, Zhou M, Dong X, et al. Epidemiological and clinical characteristics of 99 cases of 2019 novel coronavirus pneumonia in Wuhan, China: a descriptive study. Lancet 2020; 395(10223): 507–513

6. Munster VJ, Koopmans M, van Doremalen N, van Riel D, de Wit E. A novel coronavirus emerging in China—key questions for impact assessment. N Engl J Med 2020;382(8): 692–694.

7. Perlman S. Another decade, another coronavirus. N Engl J Med 2020; 382:760–762.

8. Paules CI, Marston HD, Fauci AS. Coronavirus infections—more than just the common cold. JAMA 2020; 323(8): 707–708.

9. World Health Organization. Novel Coronavirus (2019-nCoV) situation reports. https://www.who.int/docs/default-source/coronaviruse/situation-reports/20200318-sitrep-58-covid-19.pdf?sfvrsn=20876712_2 (assessed March 18th, 2020).

10. World Health Organization. Rolling updates on coronavirus disease (COVID-19). https://www.who.int/emergencies/diseases/novel-coronavirus-2019/events-as-they-happen (assessed March 11th, 2020).

11. Fehr A R, Perlman S. Coronaviruses: an overview of their replication and pathogenesis. Coronaviruses. Humana Press, New York, NY, 2015: 1–23.

12. Lee N, Hui D, Wu A, et al. A major outbreak of severe acute respiratory syndrome in Hong Kong. N Engl J Med 2003; 348:1986–94.

13. Peiris J, Lai S, Poon L, et al. Coronavirus as a possible cause of severe acute respiratory syndrome. Lancet 2003; 361:1319–25.

14. Zaki AM, Van Boheemen S, Bestebroer TM, Osterhaus AD, Fouchier RA. Isolation of a novel coronavirus from a man with pneumonia in Saudi Arabia. N Engl J Med 2012; 367:1814–20.

15. Zumla A, Hui DS, Perlman S. Middle East respiratory syndrome. Lancet 2015; 386:995–1007.

16. de Wit E, van Doremalen N, Falzarano D, Munster VJ. SARS and MERS: recent insights into emerging coronaviruses. Nat Rev Microbiol 2016;14:523.

17. World Health Organization. Severe acute respiratory syndrome - SARS. https://www.who.int/wer/2003/en/wer7835.pdf?ua=1 (assessed August 29th, 2003).

18. World Health Organization. Middle East respiratory syndrome coronavirus (MERS-CoV). https://www.who.int/emergencies/mers-cov/en/ (assessed August 2th, 2019).

19. Tsang KW, Ho PL, Ooi GC, et al. A cluster of cases of severe acute respiratory syndrome in Hong Kong. N Engl J Med 2003; 348:1977–85.

20. Shen Z, Ning F, Zhou W, et al. Superspreading sars events, Beijing, 2003. Emerging Infect Dis 2004; 10:256.

21. Cho SY, Kang J-M, Ha YE, et al. MERS-CoV outbreak following a single patient exposure in an emergency room in South Korea: an epidemiological outbreak study. Lancet 2016; 388:994–1001.

22. Kang CK, Song K-H, Choe PG, et al. Clinical and epidemiologic characteristics of spreaders of Middle East respiratory syndrome coronavirus during the 2015 outbreak in Korea. J Korean Med Sci 2017;32:744–9.

23. Chan JF-W, Yuan S, Kok K-H, et al. A familial cluster of pneumonia associated with the 2019 novel coronavirus indicating person-to-person transmission: a study of a family cluster. Lancet 2020; 395(10223): 514–523.

24. Phan LT, Nguyen TV, Luong QC, et al. Importation and human-to-human transmission of a novel coronavirus in Vietnam. N Engl J Med 2020; 382(9): 872–874.

25. Rothe C, Schunk M, Sothmann P, et al. Transmission of 2019-nCoV infection from an asymptomatic contact in Germany. N Engl J Med 2020; 382:970–971

26. Coronavirus disease (COVID-19) technical guidance: Surveillance and case definitions. https://www.who.int/internal-publications-detail/considerations-in-the-investigation-of-cases-and-clusters-of-covid-19 (accessed March 13th, 2020).

27. New coronavirus pneumonia prevention and control program (7nd ed.) (in Chinese). 2020 http://www.nhc.gov.cn/yzygj/s7653p/202003/46c9294a7dfe4cef80dc7f5912eb1989.shtml. (accessed March 4th, 2020).

28. Laboratory diagnostics for novel coronavirus. WHO 2020. https://www.who.int/health-topics/coronavirus/laboratory-diagnostics-for-novel-coronavirus (accessed March 13th, 2020).

29. World Health Organization. Laboratory testing for 2019 novel coronavirus (2019-nCoV) in suspected human cases Interim guidance. 2020 https://www.who.int/publications-detail/laboratory-testing-for-2019-novel-coronavirus-in-suspected-human-cases-20200117 (accessed March 2th, 2020).

30. Wang D, Hu B, Hu C, et al. Clinical characteristics of 138 hospitalized patients with 2019 novel coronavirus–infected pneumonia in Wuhan, China. JAMA 2020; 323(11):1061–1069.

